## Supplementary file for "SEROLOGICAL TESTING OF BLOOD DONORS TO CHARACTERISE THE IMPACT OF COVID-19 IN MELBOURNE, AUSTRALIA, 2020"

**S1 Table. List of postcodes, by sampling group**

| Low incidence <3 cases/1,000 population | Medium incidence 3-7 cases/1,000 population | High incidence >7 cases/1,000 population |
|---|---|---|
| 3071 | 3006 | 3003 |
| 3088 | 3012 | 3004 |
| 3103 | 3013 | 3011 |
| 3105 | 3028 | 3019 |
| 3106 | 3036 | 3020 |
| 3115 | 3039 | 3021 |
| 3116 | 3042 | 3022 |
| 3125 | 3048 | 3023 |
| 3126 | 3054 | 3024 |
| 3127 | 3055 | 3025 |
| 3129 | 3056 | 3026 |
| 3131 | 3057 | 3027 |
| 3133 | 3059 | 3029 |
| 3135 | 3068 | 3031 |
| 3142 | 3072 | 3034 |
| 3146 | 3093 | 3037 |
| 3153 | 3143 | 3046 |
| 3154 | 3175 | 3047 |
| 3161 | 3177 | 3051 |
| 3162 | 3181 | 3052 |
| 3167 | 3186 | 3060 |
| 3173 | 3335 | 3061 |
| 3178 | 3805 | 3064 |
| 3183 | 3809 | 3065 |
| 3184 | 3975 | 3066 |
| 3193 | 3976 | 3074 |
| 3197 | 3977 | 3076 |
| 3198 | | 3081 |
| 3201 | | 3090 |
| 3204 | | 3336 |
| 3205 | | 3337 |
| 3428 | | 3338 |
| 3800 | | 3427 |
| 3807 | | 3750 |
| 3911 | | 3753 |
| | | 3803 |
| | | 3978 |

### S1 File: Multilevel regression and poststratification modelling

The following multilevel model, including main effects for postcode, sampling stratum, sex, age and socioeconomic quintile, was specified for $\pi_i$, the true probability of SARS-CoV-2 total antibody present for specimen $i$:

$$\pi_i = \text{logit}^{-1}\left(\beta_1 + \beta_2 \text{male}_i + \beta_3 \text{SES}_i + \alpha^{\text{age}}_{\text{age}[i]} + \alpha^{\text{SES}}_{\text{SES}[i]} + \alpha^{\text{pcode}}_{\text{pcode}[i]} + \alpha^{\text{strata}}_{\text{strata}[i]}\right),$$

where, male is a variable that takes on the value of 0 for women and 1 for men, SES represents socioeconomic quintiles measured at the postcode-level, age[i], SES[i], sex[i], pcode[i] and strata[i] are index variables for specimen i, $\beta_1$, $\beta_2$ and $\beta_3$ are logistic regression coefficients and the α parameters are vectors of varying coefficients (random effects). These coefficients have hierarchical priors as follows:

$$\alpha^{\text{age}} \sim \text{normal}(0, \sigma^{\text{age}})$$

$$\alpha^{\text{SES}} \sim \text{normal}\left(0, \sigma^{\text{SES}}\right)$$

$$\alpha^{\text{pcode}} \sim \text{normal}\left(0, \sigma^{\text{pcode}}\right)$$

$$\alpha^{\text{strata}} \sim \text{normal}(0, \sigma^{\text{strata}})$$

Prior distributions for model parameters were specified as per Gelman & Carpenter (2020). That is, $\sigma^{\text{age}}$, $\sigma^{\text{SES}}$, $\sigma^{\text{pcode}}$, $\sigma^{\text{strata}}$ were assigned $\text{normal}^{+}(0,0.5)$ priors, which allows the prevalence to vary moderately by these covariates. In the primary analysis, a unit logistic prior was defined for the centred intercept $\beta_1 + \beta_2 \overline{\text{male}} + \beta_3 \overline{\text{SES}}$ (where $\overline{\text{male}}$ and $\overline{\text{SES}}$ represent the proportion male and the mean SES in the sample, respectively). This corresponds to a flat uniform (0, 1) prior distribution for the probability that an average person in the sample has the antibody. In sensitivity analysis, an alternative logistic (–3.5,1) prior distribution was specified, which focuses on values for seroprevalence below 5% (prior probability prevalence less than 2% and less than 5% were 0.40 and 0.64 respectively).

The following expression defines the observed prevalence $p$, given the true prevalence $(\pi)$ and test sensitivity $(\delta)$ and specificity $(\gamma)$:

$$p = \pi\delta + (1 - \pi)(1 - \gamma).$$

Poststratification adjustment was used to produce an estimate of prevalence for the Melbourne metropolitan blood donor population from 2019 ($N$ = 29,731; see Table 1 for summary of basic demographics) as well as the Melbourne metropolitan resident population aged 20–69 years ($N$ = 2,678,532; Table 1)

**S2 Table. Crude and estimated SARS-CoV-2 seroprevalence and 90% credible intervals (CrI) for the Melbourne blood donor population (A) and metropolitan Melbourne resident population (B) aged 20–69 years**

| | Crude estimates N (%) | A. Melbourne blood donor population | | B. Melbourne resident population | |
|---|---|---|---|---|---|
| | | Primary analysis[a] % (90% CrI) | Sensitivity analysis[b] % (90% CrI) | Primary analysis[a] % (90% CrI) | Sensitivity analysis[b] % (90% CrI) |
| **Overall population** | 77 (1.60) | 0.87 (0.25–1.49) | 0.79 (0.20–1.43) | 0.90 (0.26–1.51) | 0.82 (0.21–1.46) |
| **Sampling stratum[c]** | | | | | |
| Low incidence | 20 (1.25) | 0.73 (0.17–1.40) | 0.66 (0.13–1.33) | 0.73 (0.17–1.38) | 0.65 (0.13–1.32) |
| Medium incidence | 29 (1.81) | 0.97 (0.25–1.73) | 0.88 (0.19–1.68) | 0.99 (0.26–1.77) | 0.91 (0.20–1.71) |
| High incidence | 28 (1.75) | 1.06 (0.27–1.82) | 0.98 (0.20–1.76) | 1.06 (0.27–1.85) | 0.98 (0.20–1.79) |
| **Sex** | | | | | |
| Female | 35 (1.46) | 0.78 (0.22–1.42) | 0.71 (0.17–1.36) | 0.80 (0.23–1.44) | 0.73 (0.18–1.38) |
| Male | 42 (1.75) | 0.94 (0.25–1.68) | 0.85 (0.19–1.59) | 0.98 (0.26–1.72) | 0.89 (0.20–1.64) |
| **Age group** | | | | | |
| 20–29 years | 25 (1.92) | 0.94 (0.24–1.72) | 0.86 (0.19–1.64) | 1.00 (0.26–1.79) | 0.91 (0.20–1.72) |
| 30–39 years | 21 (1.60) | 0.88 (0.22–1.58) | 0.80 (0.18–1.52) | 0.89 (0.23–1.59) | 0.82 (0.18–1.53) |
| 40–49 years | 10 (1.09) | 0.73 (0.17–1.41) | 0.66 (0.14–1.36) | 0.75 (0.17–1.42) | 0.67 (0.14–1.37) |
| 50–59 years | 11 (1.45) | 0.79 (0.18–1.50) | 0.72 (0.15–1.43) | 0.84 (0.20–1.55) | 0.76 (0.16–1.48) |
| 60–69 years | 10 (1.95) | 0.86 (0.20–1.71) | 0.78 (0.16–1.64) | 0.91 (0.22–1.76) | 0.83 (0.17–1.69) |
| **Socioeconomic quintiles[d]** | | | | | |
| Quintile 1 (lowest) | 13 (1.71) | 1.13 (0.25–2.15) | 1.04 (0.18–2.09) | 1.11 (0.25–2.10) | 1.03 (0.19–2.04) |
| Quintile 2 | 18 (1.95) | 1.11 (0.28–1.95) | 1.02 (0.20–1.90) | 1.10 (0.28–1.94) | 1.02 (0.20–1.90) |
| Quintile 3 | 14 (1.86) | 0.98 (0.24–1.84) | 0.90 (0.18–1.77) | 0.99 (0.24–1.85) | 0.91 (0.18–1.78) |
| Quintile 4 | 10 (1.34) | 0.77 (0.17–1.49) | 0.69 (0.13–1.43) | 0.75 (0.17–1.46) | 0.68 (0.13–1.41) |
| Quintile 5 (highest) | 22 (1.38) | 0.69 (0.14–1.39) | 0.62 (0.11–1.32) | 0.68 (0.14–1.37) | 0.61 (0.11–1.30) |

[a]Estimation assumed a uniform prior distribution for seroprevalence.
[b]Estimation assumed an alternative prior distribution, which focused on values for seroprevalence below 5%.
[c]Low incidence postcodes: <3 cases/1,000 population; Medium incidence postcodes: 3-7 cases/1,000 population; High incidence postcodes: >7 cases/1,000 population (S1 Table).
[d]Assigned from residential postcode based on ABS 2016 Index of relative socioeconomic disadvantage ranking within Victoria.
